# Evaluation of *Mycobacterium tuberculosis*-specific IFN-γ, TNF-α, CXCL10, IL2, CCL2, CCL7 and CCL4 levels for active tuberculosis diagnosis

**DOI:** 10.1101/2021.11.26.21266428

**Authors:** Anastasia Fries, Vinayak Mandagere, Robert Parker, Mica Tolosa-Wright, Luis C. Berrocal-Almanza, Long Hoang, Aime Boakye, Alice Halliday, Ajit Lalvani

**Author notes:** Corresponding Author (AF).

## Abstract

Our study evaluates seven previously reported biomarkers for active tuberculosis (ATB) diagnosis. We compared *Mycobacterium tuberculosis*-specific IFN-γ, TNF-α, CXCL10, IL2, CCL2, CCL7 and CCL4 levels in patients with ATB and non-tuberculosis respiratory diseases. Our ATB group included equal numbers of patients with positive and negative QuantiFERON-TB Gold In-Tube (QFT-GIT) results, to assess whether any biomarker offered superior diagnostic accuracy to IFN-γ. No biomarker achieved higher sensitivity than QFT-GIT for ATB diagnosis without significant loss of specificity. Our study design provides an efficient strategy for rapidly gating future biomarkers by using clinically relevant and representative patient groups in whom current QFT-GIT tests fail.

## Background

Tuberculosis (TB) remains one of the most devasting infectious diseases, with approximately 10 million incident cases and 1.2 million deaths in 2019[1]. New, innovative diagnostic tests are urgently needed to facilitate prompt diagnosis, effective treatment and prevent disease transmission. Two major unmet clinical needs remain for active TB (ATB) diagnosis[2]. The first is a rapid, accurate test to identify TB in culture-negative cases; the second is a simple point-of-care, or *rule-out* test.

The ‘Interferon gamma release assays for Diagnostic Evaluation of Active TB ’ (IDEA) Study is one of the largest prospective cohort studies to investigate the clinical utility of Interferon Gamma Release Assays (IGRAs) for ATB diagnosis in a low-incidence setting[3]. IDEA provided substantial, generalisable, high-quality evidence that existing IGRAs do not have a useful role as rule-out tests for ATB owing to their insufficient diagnostic sensitivity, 81.4% for T-SPOT.TB and 67.3% for QFT-GIT.

Recent biomarker discovery platforms including high-throughput gene-expression by RNA-Seq, proteomics and flow-cytometry have identified alternative biomarkers to IFN-γ, which might provide greater sensitivity for ATB diagnosis and offer rapid, rule-out tests for ATB[4]. Several studies reported promising results regarding the diagnostic potential of *Mycobacterium tuberculosis (Mtb)*-specific chemokines and cytokines[5][6][7][8]. However, these studies mostly lack clinically relevant control populations, using healthy controls or healthy persons with latent TB infection (LTBI), instead of patients with other diseases that clinically mimic TB, often with concomitant LTBI. Some studies also excluded individuals lacking culture-confirmation or other difficult-to-diagnose but important and common TB subgroups, such as extrapulmonary TB, in whom new diagnostic tests would be most beneficial.

We aimed to efficiently assess whether seven previously identified, promising biomarkers (IFN-γ, TNF-α, CXCL10, IL2, CCL2, CCL7 and CCL4) could distinguish ATB patients within a cohort of patients presenting with the full clinical spectrum of suspected TB in routine practice. We designed a nested case-control study within IDEA, to accurately assess each biomarker’s performance compared to IFN-γ for ATB diagnosis. Uniquely, for the current study we enriched our ATB study population for patients in whom current IGRAs fail, i.e. ATB cases with false-negative IGRA-negative results. These are the cases where the unmet need is greatest and they provide an opportunity rapidly to assess whether candidate biomarkers can offer increased sensitivity compared to IFN-γ measured by IGRA.

We exploited the unique value of the IDEA cohort to evaluate the real-life diagnostic performance of *Mtb*-specific biomarkers at distinguishing ATB from its naturally-occurring differential diagnoses. We used a ‘gating’ approach for biomarker evaluation, initially evaluating their diagnostic accuracy in ∼10% of the IDEA cohort, to establish ‘proof-of-principle’ that biomarkers could achieve an increase in sensitivity for ATB diagnosis, without significantly compromising specificity. Only biomarkers meeting this criterion would then be further assessed in the full IDEA cohort.

## Methods

### Study group selection

Our study was nested within the large, prospective, multi-centre IDEA study, which recruited adults with suspected ATB at the point of diagnostic work-up in routine clinical practice in England[3]. The Study Flow Chart (**Fig 1**) shows the derivation of our case-control study from the full IDEA cohort. IDEA was purpose-designed for the rigorous assessment of novel ATB diagnostics tests in low-incidence settings. Participants provided written, informed consent. The study was approved by Camden and Islington National Research Ethics Committee (reference 11/H0722/8/).

**Fig 1.**
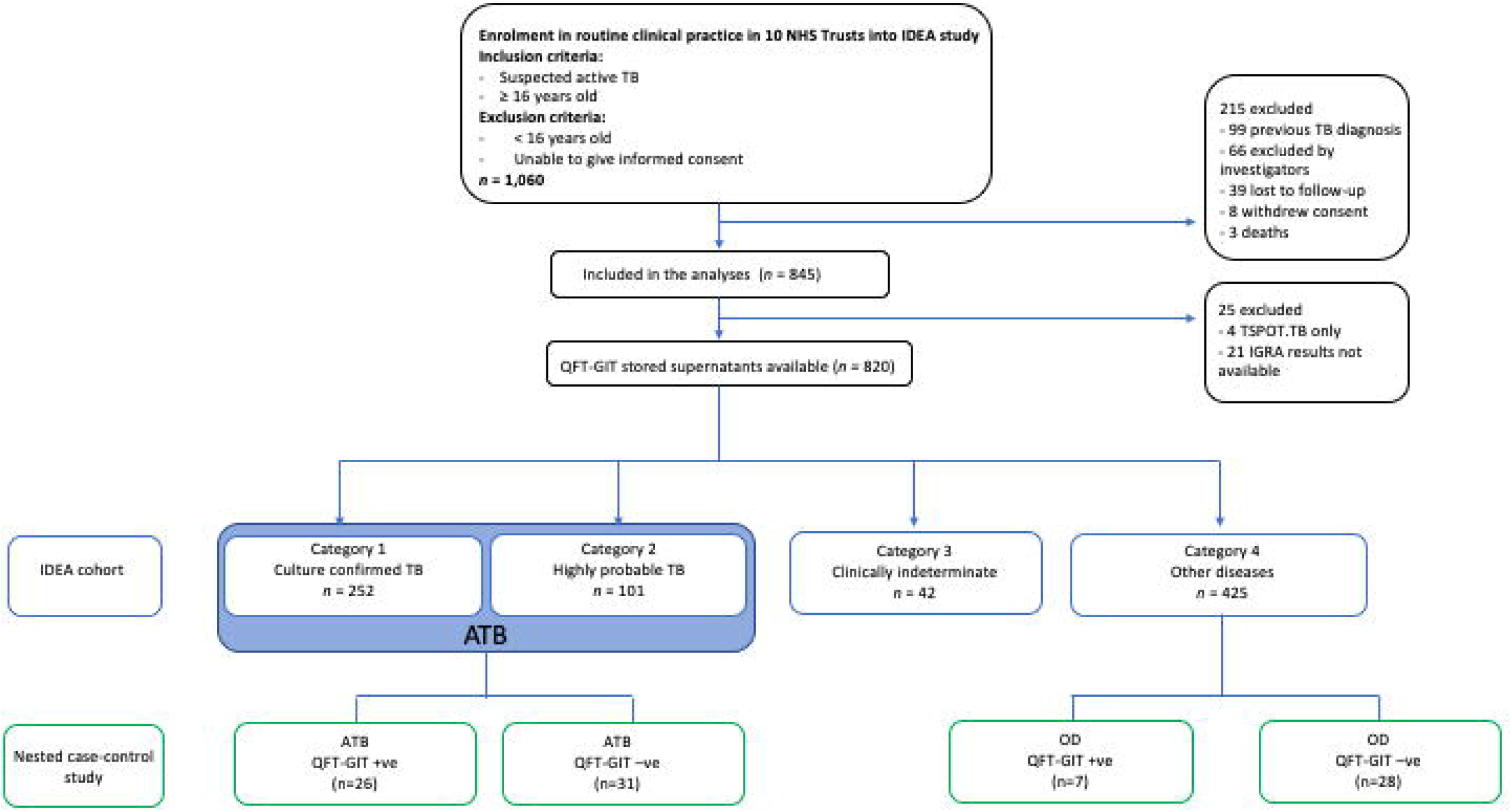
Study design and sample selection within the IDEA study cohort. Patients were prospectively recruited to the IDEA study from routine clinical practice at the point of clinical suspicion of TB and diagnostic work-up and were followed up for 12 months. With all available clinical, radiological, microbiological and histological data participants received a definitive diagnosis and were classified as Category 1 (culture-confirmed TB); Category 2 (highly-probable TB); Category 3 (clinically indeterminate); Category 4 (other diseases). Categories 1 and 2 comprise all ATB. In our nested case-control study, patients in category 3 were excluded (n= 42) as were those who withdrew consent or were lost to follow up (n= 215). Of the remaining patients eligible for analysis (n = 845), QFT-GIT results were available for n = 820. We used random computer-generation to select 30 patients within the three main categories (ATB, QFN-GIT positive; ATB QFT-GIT negative; OD QFT-GIT negative) and 10 patients from within OD QFT-GIT positive. Of these, three samples could not be located and five had insufficient volume leaving a total of 92 patients.

In IDEA, pre-treatment blood samples from each study participant were tested with all IGRAs and definitive diagnostic categorisation verified according to stringent, pre-defined, previously validated criteria (Categories 1 – 4)[9]. Category 1 (culture-confirmed TB): suggestive clinical and radiological findings AND positive *Mtb* culture. Category 2 (highly-probable TB): clinical and radiological features highly suggestive of TB AND no positive culture AND clinician decision-to-treat AND appropriate response to therapy AND supportive histological evidence if available. Category 3 (clinically indeterminate): TB was neither highly-probable nor reliably excluded. Category 4 (other diseases (OD)): no evidence of ATB using combinations of symptoms, risk factors, negative bacteriology and evidence of another diagnosis if available.

We utilised samples biobanked at enrolment into the IDEA biobank and purposively enriched for ATB patients with false-negative IGRA results. In the IDEA cohort approximately 30% of ATB patients had false-negative QFT-GIT results. Our ATB cohort included 50% such IGRA-negative ATB cases to efficiently assess each biomarker’s ability to detect ATB where IFN-γ fails to do so. We selected approximately equal numbers of IGRA-positive ATB, IGRA-negative ATB and IGRA-negative OD cases (∼30 patients per group) and 10 OD IGRA-positive patients.

In our Phase I, we analysed seven biomarkers (IFN-γ, TNF-α, CXCL10, IL2, CCL2, CCL7 and CCL4) in 32 patients comprising 11 IGRA-positive ATB cases, 5 IGRA-negative ATB cases, 7 IGRA-positive cases with OD and 9 IGRA-negative OD cases). Based on preliminary results from this small Phase I study, we analysed IFN-γ, CXCL10 and CCL2 in a further 60 patients (15 ATB IGRA-positives; 26 ATB IGRA-negatives; 19 OD IGRA-negatives) in Phase II.

### Candidate biomarker detection from QFT-GIT supernatants

We utilised the bio-repository of stored supernatants from QFT-GIT tests performed in IDEA[3]. Briefly, as described in Whitworth et al.,[3] blood was collected in QFT-GIT blood collection tubes and analysed in real time at the Tuberculosis Research Centre (Imperial College London, London, UK), according to the manufacturer’s instructions. QFT-GIT supernatants were subsequently stored at -80°C.

We analysed cytokine and chemokine concentrations using Meso Scale Discovery (MSD) U-PLEX assay platforms, as per manufacturer’s instructions. Samples were thawed at room temperature and diluted 1:2 for IFN-γ, TNF-α and IL2 and 1:40 for CXCL10, CCL2, CCL7 and CCL4. For each sample, mitogen-stimulated, antigen-stimulated and nil-stimulated readings were analysed. Duplicate readings were averaged for antigen-stimulated samples. Plates were read using MESO QuickPlex SQ 120 instrument. Biomarker concentrations were calculated by normalising the antigen-stimulated from nil values and expressed in pg/ml.

### Statistical analysis

All statistical analysis was performed using GraphPad Prism version 8.0. We analysed differences in normalised biomarker concentrations between groups using Kruskal-Wallis tests. ROC curve analyses determined cut-off values to achieve maximum sensitivity for minimal loss of specificity.

## Results

### Study cohort

A total of 1,060 participants with suspected TB were enrolled into IDEA, of which 92 were included in our study (**Fig 1**). Participant’s demographic and clinical characteristics and diagnostic categorisations are summarised in **S1 and S2 Tables**. The QFT-GIT sensitivity and specificity values in our study population were 45.6% and 80% respectively.

### Phase I. Preliminary screening of Mtb-specific IFN-γ, TNF-α, CXCL10, IL2, CCL2, CCL7 and CCL4 biomarkers for ATB diagnosis

The diagnostic performance of each of the 7 biomarker was determined by its ability to detect true-positive (TP) and true-negative (TN) results (**S3 Table**).

MSD-measured biomarkers (except CCL4) detected higher numbers of TP compared to QFT-GIT, however, all biomarkers lost specificity, with a 42.9% increase in false-positive (FP) results. MSD-measured IFN-*γ* detected more TP in ATB patients (15 TP) compared to QFT-GIT-measured IFN-*γ* (11 TP). After IFN-*γ*, CXCL10 detected the next highest number of TPs in the ATB group (14 TP). CCL4 performed inferiorly to QFT-GIT, with increased false-negative (FN) and FP results.

TNF-*α* and IL2 had low biomarker concentrations, therefore, these were excluded from further analysis. Although CCL7 detected one more TP than CCL2, CCL7 also had very low concentrations, so we decided to only analyse CCL7 if CCL2 demonstrated promise as a biomarker for ATB diagnosis in Phase II. Based on these results, we proceeded to analyse IFN-γ, CXCL10 and CCL2 in Phase II.

### Phase II. Further evaluation of Mtb-specific IFN-γ, CXCL10 and CCL2 biomarker analysis for ATB diagnosis

In Phase II, we analysed three biomarkers (IFN-γ, CXCL10 and CCL2) in a further 60 patients, resulting in a total of 92 patients (**Table 1**). CXCL10 achieved the highest increase in TP results for ATB diagnosis, with 43 TP compared with 26 TP results for QFT-GIT. MSD-measured IFN-*γ* detected 41 TP results, while CCL2 only detected the same number of TP results as QFT-GIT (26 TP). All three biomarkers demonstrated >35% loss of specificity compared to QFT-GIT, giving 19-20 FP results compared to only 7 FP results with QFT-GIT.

**Table 1.**
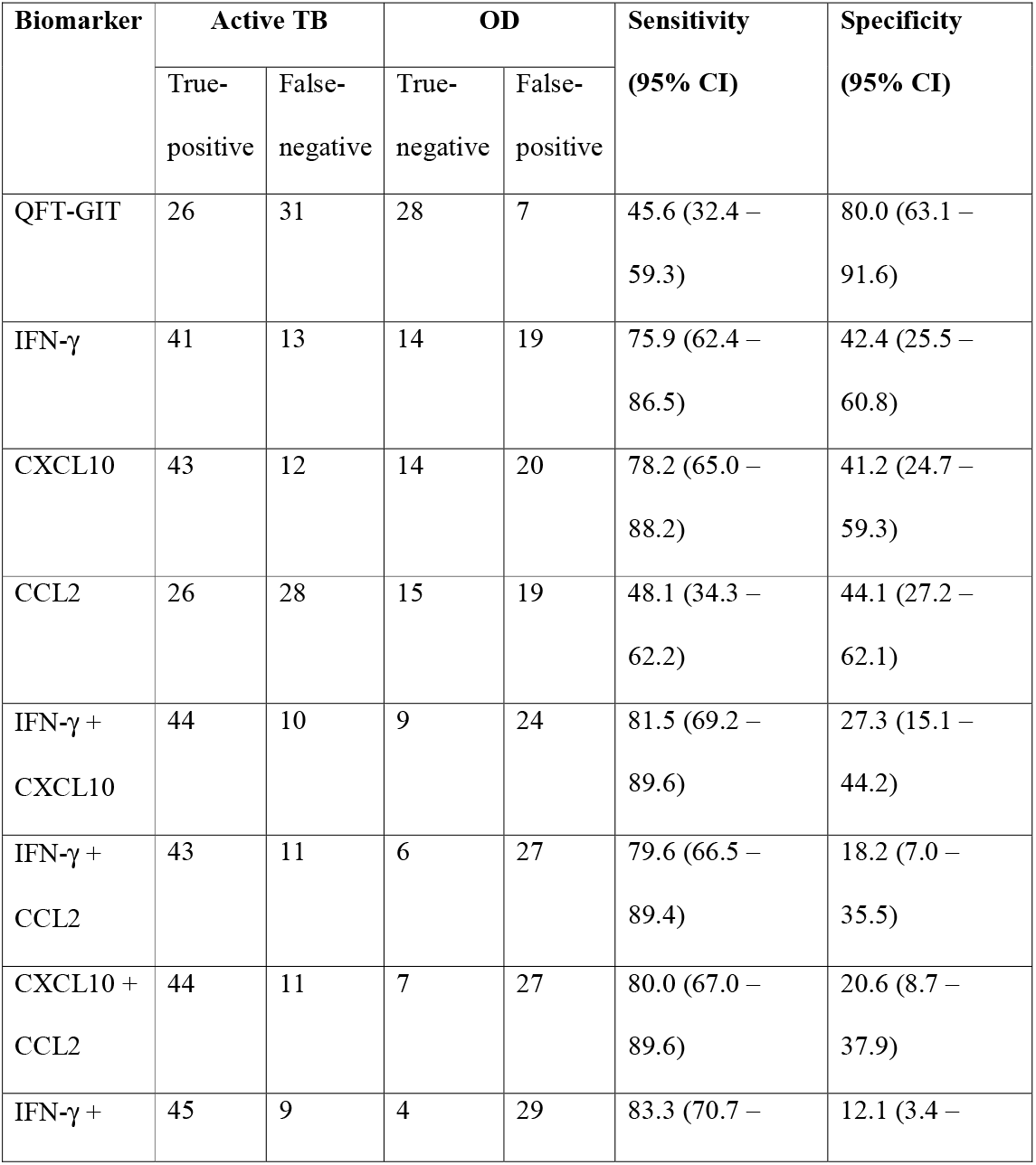
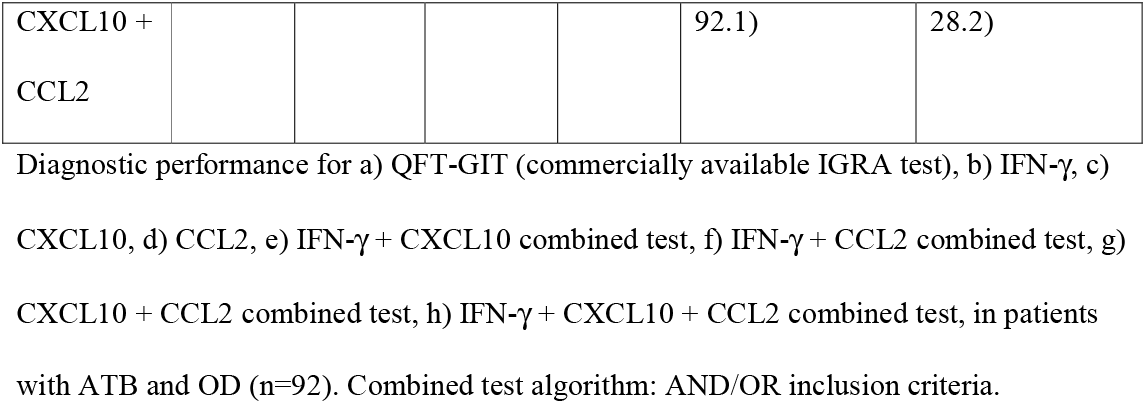
Biomarker performance of *Mtb*-specific IFN-γ, CXCL10 and CCL2 for ATB diagnosis.

Combined dual biomarker analyses in a “Yes AND/OR Yes” test achieved sensitivity values of between 79.6% - 81.5% but specificity decreased to 18.2% - 27.3%. A ‘triple-test’ combining IFN-γ, CXCL10 and CCL2 results achieved the highest number of TP results (45) with a sensitivity of 83.3% and specificity of 12.1%.

## Discussion

To our knowledge, this is the first case-control study to enrich ATB cases for IGRA-negative ATB cases from an observational cohort of patients with suspected TB presenting in routine clinical practice. This enabled us efficiently to assess the diagnostic performance of promising biomarkers for ATB diagnosis in the exact population who are currently misdiagnosed using available immune-based tests, i.e. those in whom the clinical need is greatest. We assessed in a head-to-head comparison whether any of 7 promising biomarkers were superior to IFN-γ in their ability to discriminate patients with ATB by using equal numbers of IGRA-positive and IGRA-negative ATB cases.

No biomarker achieved sufficient sensitivity for ATB diagnosis without unacceptably compromised specificity (Phase I: >40% increase in OD FPs; Phase II >170% increase in OD FPs). The high negative predictive value (NPV) of QFT-GIT for LTBI means IGRA-negative OD controls are very likely TN results[10]. Consequently, the substantial number of positive results for each of the biomarkers among the QFN-GIT negative, TB unexposed OD case are very likely FP results. Due to low biomarker specificities, we did not proceed with further analysis in the full IDEA cohort.

The WHO target product profile (TPP) states that TB triage tests for individuals requiring further confirmatory testing should reach minimum 90% sensitivity and 70% specificity (optimal criteria: 95% sensitivity and 80% specificity)[2]. Currently-available QFT-GIT tests provide a sensitivity of ∼67% for active TB and a growing body of evidence suggests multiple markers (biosignatures) will be required to reach TPP-level sensitivity and triage test performance[11][12]. This is unsurprising given the complex immune responses in TB and the fact that diseases other than TB affect circulating biomarker concentrations.

Our ‘triple-test’ (IFN-γ, CXCL10 and CCL2) achieved 83.3% sensitivity, which is promising in a study population enriched to include 50% ATB cases with false-negative IGRA results. However, this level of sensitivity was accompanied by a specificity of only 12.1%, which falls well below the WHO TPP requirements. Although a triage-test with ∼15% specificity would result in an unacceptably high proportion of patients requiring further screening tests, the test’s NPV might increase in lower prevalence populations with lower pre-test probabilities of TB.

An analogous example from clinical practice is the use of serum D-dimer tests to exclude venous thromboembolic disease in patients with low pre-test probability (low-risk Wells Score)[13]. Serum D-dimers have a sensitivity >95%[14] and specificity ∼40-60%, which varies depending on clinical context[15]. An equivalent ATB test could be useful in cohorts with very low pre-test probability, for example screening of patients awaiting initiation of biological therapies or transplantation, in whom exclusion of TB prior to treatment is essential. Such patient groups would have a lower prevalence of TB than our cohort of suspected TB patients, therefore the NPV of a rule-out test for ATB may be higher and provide a useful, prompt rule-out test. Notwithstanding, the 12% specificity of the sensitive combination of IFN-γ, CXCL10 and CCL2 makes the likelihood of developing a clinically useful test based on these biomarkers extremely low.

The strengths of our study include rigorous case-definition for highly-probable TB in a clinically-relevant cohort of patients with suspected TB[3]. We leveraged the cohort to create a nested case-control study purposely constituted to enable efficient gating of candidate biomarkers for ATB diagnosis. Inclusion of patients with LTBI within the OD group reflects the diagnostic uncertainty faced by clinicians treating patients with suspected TB in routine clinical practice. In low-burden countries, many patients with suspected TB are from ethnic groups with a high prevalence of LTBI and therefore more likely to be IGRA-positive when ATB is excluded.

One limitation of this study is that whilst cytokine responses were generally higher in antigen-stimulated samples, ∼14% samples had negative antigen-nil concentrations. This may be due to complications following sample freezing and ideally fresh supernatants would be used in future studies.

In summary, our study exemplifies a novel, pragmatic gating study design for efficiently assessing the diagnostic performance of candidate ATB biomarkers enabling investigators to rapidly exclude biomarkers from further evaluation.

## Data Availability

All data produced in the present study are available upon reasonable request to the authors

## Acknowledgements

We thank all the patients for their participation in the study and the medical staff and nursing at the recruiting hospitals. We are grateful to the VANTDET independent scientific advisory group and the Imperial College NIHR Health Protection Research Unit in Respiratory Infections and the Imperial College Biomedical Research Centre are also acknowledged for their support. The views expressed are those of the author(s) and not necessarily those of the NIHR or the Department of Health and Social Care.

## Authors’ Contributions

AF: Study design, Performed U-PLEX biomarker assay experiments, analysed MESO QuickPlex data, statistical analyses and wrote the manuscript. Accessed and verified the data; VM: Performed and optimised biomarker assay experiments; RP: Performed biomarker assay experiments; MT-W: Study design and accessing clinical data. LCBA: Study design and review of manuscript. LH: Study design; AB: Managed clinical data, sample sourcing; AH: Study design; statistical analyses; intellectual contribution and review of and input into manuscript; AL: Conceived, designed the study and wrote the manuscript.

