## Supplementary Tables 1, 2, 3 for "Evaluation of *Mycobacterium tuberculosis*-specific IFN-γ, TNF-α, CXCL10, IL2, CCL2, CCL7 and CCL4 levels for active tuberculosis diagnosis"

**Supporting information**

**S1 Table. Demographic and clinical characteristics of participants (*n* = 92)**

Acronyms: ATB, all active TB; QFT-GIT, QuantiFERON-TB Gold In-Tube; +, Positive; -, Negative; OD, other disease; HIV, human immunodeficiency virus; TB, tuberculosis; BCG, Bacillus Calmette–Guérin

**S2 Table. Final presenting diagnoses of patients with TB (*n* = 57) and other diseases (*n* = 35)**

Acronyms: EPTB, extrapulmonary TB; PTB, pulmonary TB; TB, tuberculosis; LRTI, lower respiratory tract infection.

**S3 Table. Biomarker performance of *Mtb*-specific IFN-γ, TNF-α, CXCL10, IL2, CCL2, CCL7 and CCL4 for ATB diagnosis**. *Diagnostic performance tables for a) QFT-GIT (commercially available IGRA test), b) IFN-γ, c) IL2, d) TNF-α, e) CXCL10, f) CCL2, g) CCL7 and h) CCL4, in patients with ATB and OD (n=32).*

**S1 Table**

| **Characteristic** | **ATB QFT-GIT+**  **(*n* = 26)** | **ATB QFT-GIT- (*n* = 31)** | **OD QFT-GIT+**  **(*n* = 7)** | **OD QFT-GIT- (*n* = 28)** |
| --- | --- | --- | --- | --- |
| ***Demographic*** |  |  |  |  |
| Median age (range), years | 34 (19 – 69) | 45 (20 – 81) | 42 (36 – 74) | 47 (18 – 84) |
| Female, n (%) | 13 (50) | 17 (54.8) | 0 (0) | 12 (42.9) |
| Ethnic origin, n (%) |  |  |  |  |
| *Asian* | 0 (0) | 4 (12.9) | 0 (0) | 2 (7.1) |
| *Black* | 3 (11.5) | 3 (9.7) | 2 (28.6) | 6 (21.4) |
| *Hispanic* | 0 (0) | 0 (0) | 0 (0) | 0 (0) |
| *Indian subcontinent* | 21 (80.8) | 18 (58.1) | 5 (71.4) | 9 (32.1) |
| *Middle Eastern* | 0 (0) | 1 (3.2) | 0 (0) | 0 (0) |
| *Mixed* | 0 (0) | 1 (3.2) | 0 (0) | 0 (0) |
| *White* | 2 (7.7) | 4 (12.9) | 0 (0) | 11 (39.3) |
| *Unknown* | 0 (0) | 0 (0) | 0 (0) | 0 (0) |
| Median BMI (range) | 25 (18 – 34) | 25 (19 – 34) | 24 (18 – 29) | 25 (16 – 43) |
| History of previous TB, n (%) | 2 (7.7) | 1 (3.2) | 0 (0) | 0 (0) |
| BCG vaccine, n (%) | 18 (69.2) | 24 (77.4) | 7 (100) | 24 (85.7) |
| Co-morbidities, n (%) |  |  |  |  |
| HIV | 0 (0) | 1 (3.2) | 0 (0) | 0 (0) |
| Asthma | 0 (0) | 0 (0) | 0 (0) | 1 (3.6) |
| Diabetes | 2 (7.7) | 7 (22.6) | 3 (42.9) | 4 (14.3) |
| Other | 4 (15.4) | 4 (12.9) | 3 (42.9) | 4 (14.3) |
| Symptoms |  |  |  |  |
| Cough, n (%) | 9 (34.6) | 12 (38.7) | 3 (42.9) | 10 (35.7) |
| Fever, n (%) | 12 (46.2) | 23 (74.2) | 4 (57.1) | 21 (75.0) |
| Night sweats, n (%) | 5 (19.2) | 13 (41.9) | 2 (28.6) | 13 (46.4) |
| Weight loss, n (%) | 12 (46.2) | 18 (58.1) | 4 (57.1) | 15 (53.6) |
| Haemoptysis, n (%) | 3 (11.5) | 4 (12.9) | 2 (28.6) | 2 (7.1) |
| Lethargy, n (%) | 12 (46.2) | 14 (45.2) | 2 (28.6) | 16 (57.1) |
| Other, n (%) | 8 | 18 (58.1) | 3 (42.9) | 13 (46.4) |

**S2 Table**

| ***n* (%)** | **All TB**  **(*n* = 55)** | **Other diseases**  **(*n* = 35)** |
| --- | --- | --- |
| Culture negative | 16 (29.1) | 28 (80.0) |
| Culture not done | 0 (0) | 7 (20.0) |
| Smear positive TB | 11 (20.0) | 0 (0) |
| Smear negative TB | 37 (67.3) | 28 (80.0) |
| Smear not tested | 7 (12.7) | 7 (20.0) |
| PTB | 20 (36.4) | n/a |
| EPTB | 31 (56.4) | n/a |
| PTB + EPTB | 4 (7.3) | n/a |
| Pneumonia | n/a | 9 (25.7) |
| Sarcoidosis | n/a | 0 (0) |
| Cancer | n/a | 3 (8.6) |
| Non-pneumonia LRTI | n/a | 4 (11.4) |
| Bronchiectasis | n/a | 0 (0) |
| Reactive lymphadenopathy | n/a | 0 (0) |
| Upper respiratory tract infection | n/a | 2 (5.7) |
| Asthma | n/a | 1 (2.9) |
| Other diagnoses | n/a | 17 (48.6) |

**S3 Table.**

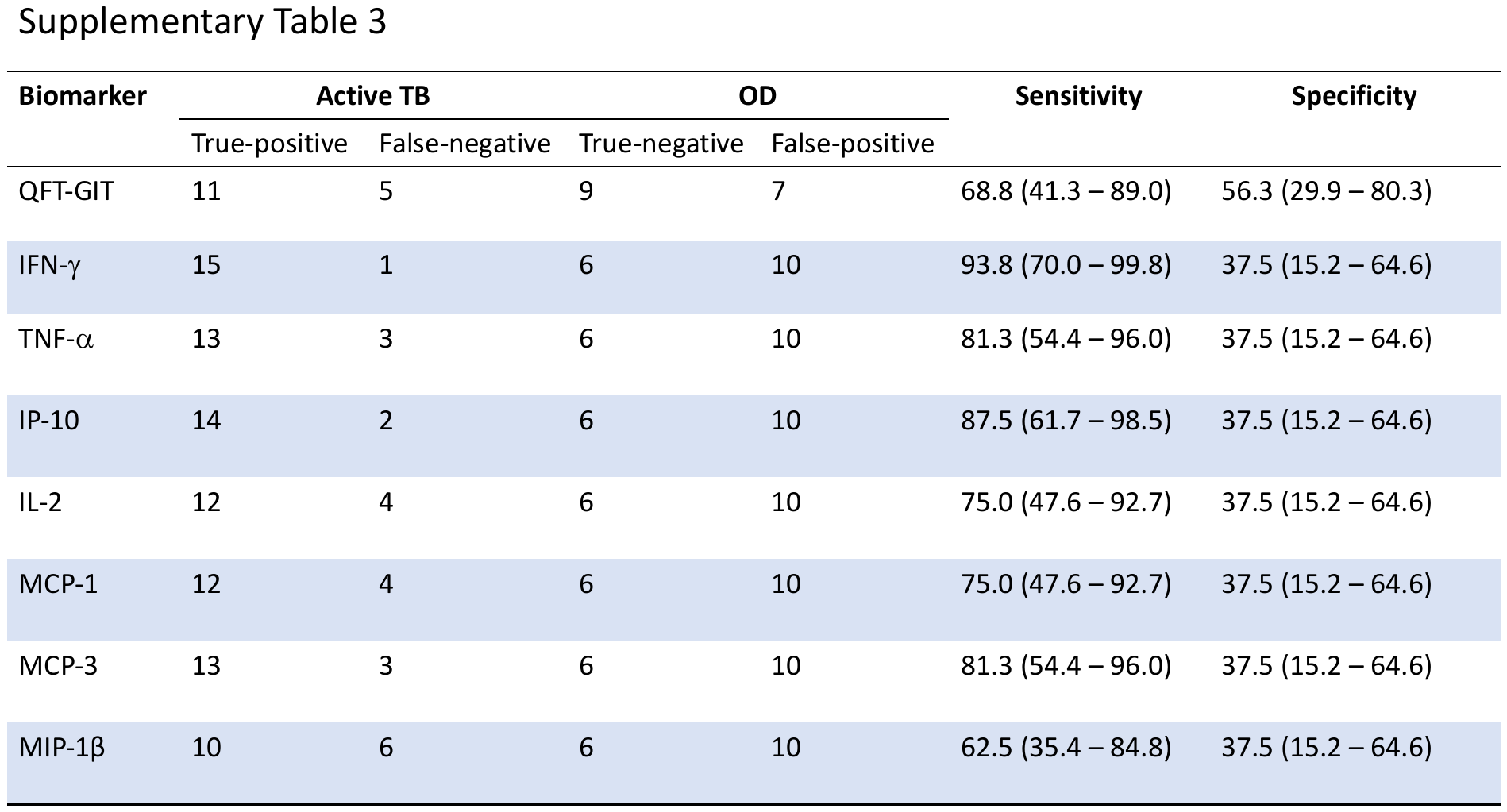
